# Establishment and Efficacy of an Endoscopic Pathogen Visualization Literacy (EPVL) Training Program for Gastroenterologists Based on Fluorescence Rapid On-Site Evaluation (ROSE) Technology

**DOI:** 10.64898/2026.08.12.26360123

**Authors:** Lin Zhang, Yanhong Hou, Binghui Li, Kai Wu, Jing Zhang, Mi Yang

## Abstract

**Objective:** To establish a standardized training program for endoscopic pathogen visualization literacy (EPVL) based on fluorescence rapid on-site evaluation (ROSE) technology for gastroenterologists, and to evaluate its training efficacy.

**Methods:** A prospective quasi-experimental study was conducted. A total of 54 gastroenterology trainees were non-randomly allocated into the EPVL training group (Group A, n=28, 16-hour comprehensive training) and the control group (Group B, n=26, 3.5-hour traditional teaching). Pre- and post-training assessments included theoretical examinations, fluorescence ROSE image interpretation tests (30 parallel images per set), interpretation speed measurement, and clinical decision-making integration evaluation. The primary outcome was the change in image interpretation accuracy, analyzed by ANCOVA with pre-test scores as the covariate.

**Results:** Baseline characteristics were comparable between groups (P>0.05 for all demographic variables and pre-test scores). Group A showed significant improvement in image interpretation accuracy from 57.8±13.6% pre-training to 82.5±11.2% post-training (improvement of 24.7%, paired t=-12.86, P<0.001), while Group B improved from 58.5±13.0% to 71.0±13.5% (improvement of 12.5%, paired t=-5.24, P<0.001). After ANCOVA adjustment for pre-test scores, the between-group difference was significant (F(1, 51)=10.95, P=0.0017, η^2^=0.177), with Cohen’s d=0.94 (large effect size). Interpretation speed in Group A (19.2±2.8 s/image) was significantly faster than in Group B (32.5±6.0 s/image, t=-10.45, P<0.001). Clinical decision-making scores were significantly higher in Group A (80.5±8.0 vs. 65.3±11.5, t=5.60, P<0.001). The Kappa agreement with the gold standard in Group A improved from 0.56±0.18 to 0.84±0.11 (t=-8.35, P<0.001). Participant satisfaction exceeded 88%.

**Conclusion:** The EPVL training program significantly improves gastroenterologists’ fluorescence ROSE image interpretation accuracy, speed, and clinical decision-making integration, providing a novel and effective standardized training paradigm for digestive endoscopy education.

## 1 Introduction

### 1.1 Background

Helicobacter pylori (H. pylori) infection is one of the most common chronic infections worldwide and is closely associated with chronic gastritis, peptic ulcer disease, gastric cancer, and mucosa-associated lymphoid tissue (MALT) lymphoma[1]. The prevalence of H. pylori infection in China is as high as 40%–60%, and eradication therapy is an important first-level preventive measure against gastric cancer. However, with the rising antibiotic resistance rate of H. pylori year by year, the eradication rate of traditional “empirical” regimens has dropped below 80%, and the demand for precision diagnosis and treatment is becoming increasingly urgent[2-7].

Fluorescence rapid on-site evaluation (ROSE) technology is a novel pathogen visualization technique developed in recent years[8]. Through specific fluorescent probe labeling, it enables real-time endoscopic observation of H. pylori morphology, viability status, coccoid transformation, and spatial distribution. Compared with traditional pathological methods such as Giemsa staining and silver staining, fluorescence ROSE technology offers advantages including real-time visualization, simplicity, and reproducibility, allowing gastroenterologists to “directly visualize” H. pylori in real time for the first time[9-13]. Figure 1 illustrates the comparative detection effects of traditional Giemsa staining, silver staining, and fluorescence ROSE technology for Helicobacter pylori.

**Figure 1.**
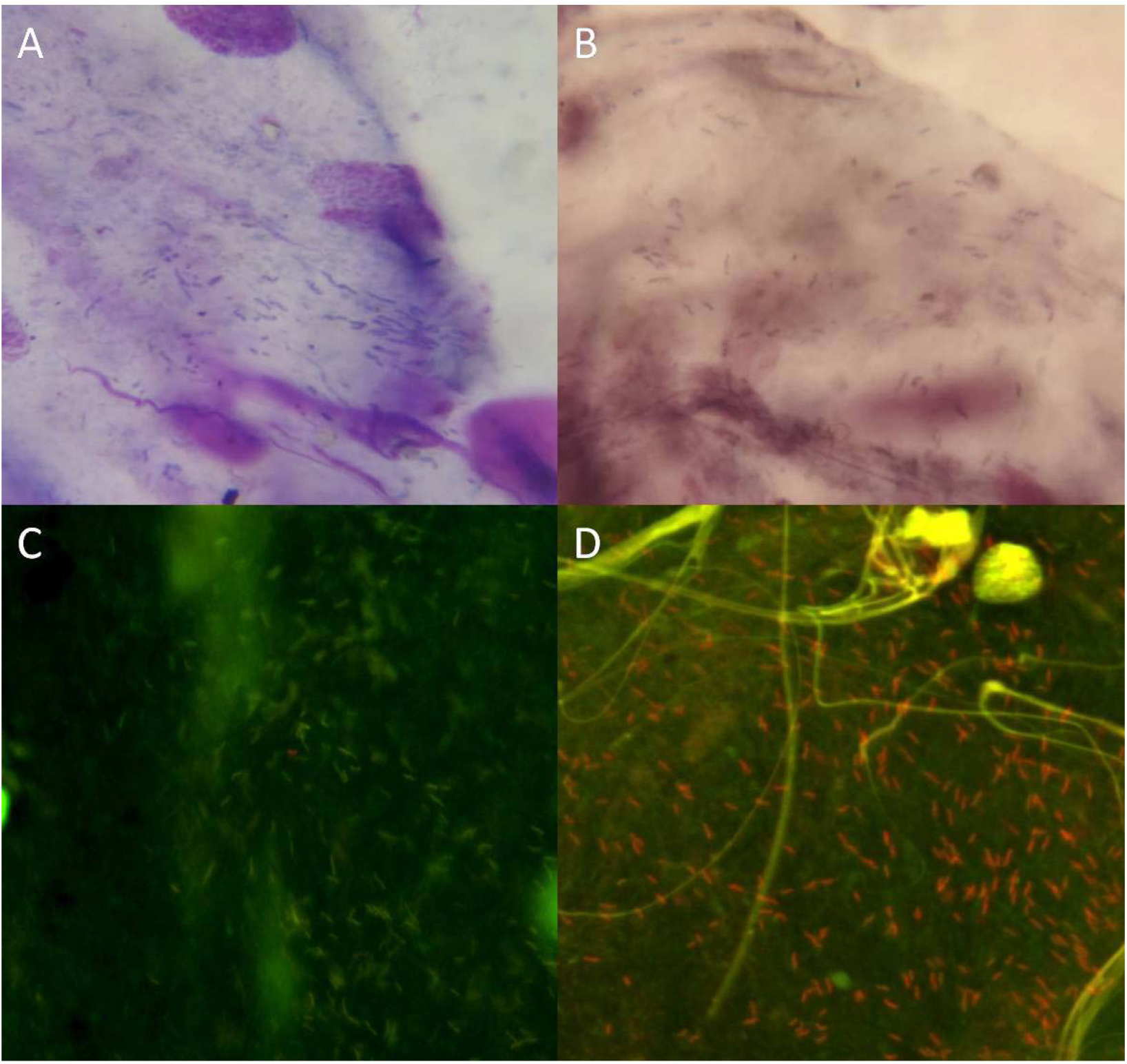
Comparison of H. pylori morphology under traditional staining and fluorescence ROSE. (A) Giemsa staining showing purple rod-shaped H. pylori (traditional pathological method). (B) Silver staining showing brown-black H. pylori (traditional pathological method). (C) Fluorescence ROSE live bacteria detection showing green fluorescent live H. pylori. (D) Fluorescence ROSE dead bacteria detection (dual staining) showing red fluorescent dead H. pylori with green mucus strands in the background. Fluorescence ROSE technology enables real-time differentiation of live and dead H. pylori during endoscopy, whereas traditional staining methods cannot distinguish bacterial viability.

However, the full clinical value of fluorescence ROSE technology depends heavily on the operator’s image interpretation competence. Currently, there is no systematic fluorescence ROSE image interpretation training program for gastroenterologists domestically or internationally[14], and most physicians lack standardized training in pathogen visualization literacy. This situation of “technology ahead, literacy lagging” seriously restricts the clinical promotion of fluorescence ROSE technology.

### 1.2 Proposal of the EPVL Concept

Based on the above background, our team first proposed the concept of “Endoscopic Pathogen Visualization Literacy” (EPVL)[15], defined as the comprehensive capability of gastroenterologists to perform real-time identification, morphological interpretation, viability assessment, and clinical decision-making integration of pathogenic microorganisms on the gastric mucosal surface using fluorescence ROSE technology during endoscopic procedures. EPVL encompasses four dimensions: (1) Knowledge dimension—principles of fluorescence ROSE technology and H. pylori pathogenesis; (2) Skill dimension—real-time image recognition and morphological interpretation ability; (3) Attitude dimension—pathogen visualization awareness and precision diagnosis/treatment philosophy; and (4) Application dimension—clinical immediate decision-making and treatment regimen adjustment capability. These four dimensions together constitute the theoretical framework of EPVL, as shown in Figure 2.

**Figure 2.**
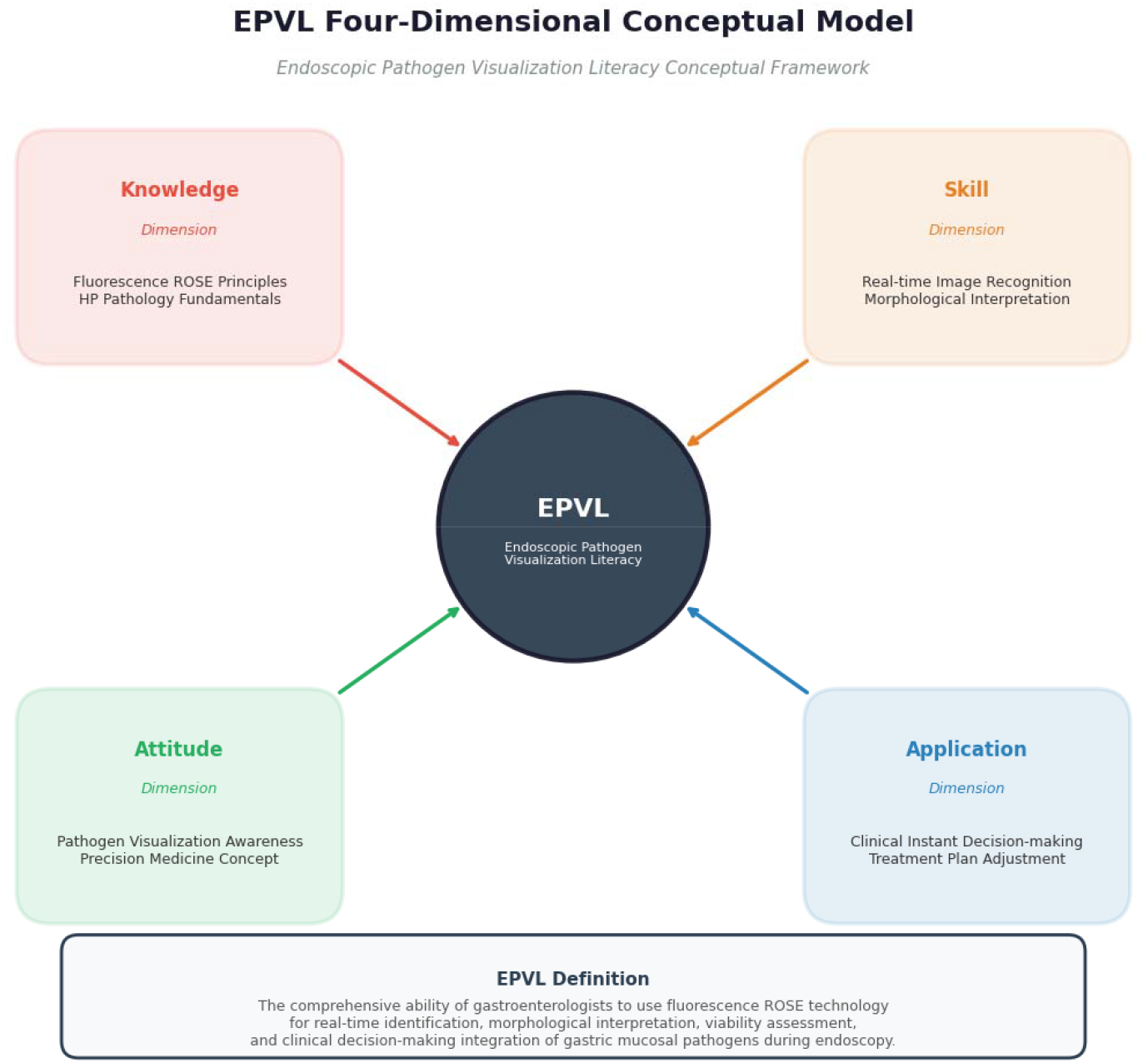
A Four-Dimensional Conceptual Framework for Endoscopic Pathogen Visualization Literacy (EPVL) EPVL comprises four core dimensions: the knowledge dimension (principles of fluorescence ROSE technology and H. pylori pathogenesis), the skill dimension (real-time image recognition and morphological interpretation ability), the attitude dimension (pathogen visualization awareness and precision diagnosis/treatment philosophy), and the application dimension (clinical immediate decision-making and treatment regimen adjustment capability). The four dimensions together constitute the comprehensive capability system for gastroenterologists to perform pathogen visualization diagnosis and treatment using fluorescence ROSE technology.

### 1.3 Research Objectives

This study aims to: (1) establish the first standardized EPVL training program in China[16]; (2) validate the effectiveness of this training program through a prospective quasi-experimental study; and (3) provide a generalizable training paradigm for digestive endoscopy specialty education.

## 2 Materials and Methods

### 2.1 Study Design

This study was a prospective quasi-experimental study (non-randomized controlled trial) employing a dual-control design of non-randomized concurrent control plus self pre-post control. The study was approved by the hospital ethics committee, and all participants signed informed consent forms.

### 2.2 Participants

Inclusion criteria: (1) practicing physicians, residents, or fellows in gastroenterology; (2) voluntary participation in this study; (3) ability to complete the entire training and assessment process.

Exclusion criteria: (1) prior experience in ROSE technology operation or interpretation; (2) interruption during the training period for any reason; (3) refusal to cooperate with assessments.

### 2.3 Group Allocation

Natural grouping by training batch: the first batch of trainees was assigned to Group A (EPVL training group), and the second batch to Group B (control group). Group A received a 16-hour comprehensive EPVL training, while Group B received 3.5 hours of traditional teaching (distribution of written materials plus a single theoretical lecture). Both groups met the hospital training curriculum requirements, and trainees were explicitly informed that this was part of the normal teaching arrangement.

### 2.4 EPVL Training Program (Group A)

The training program comprised six modules(Table 1):

**Table 1.**
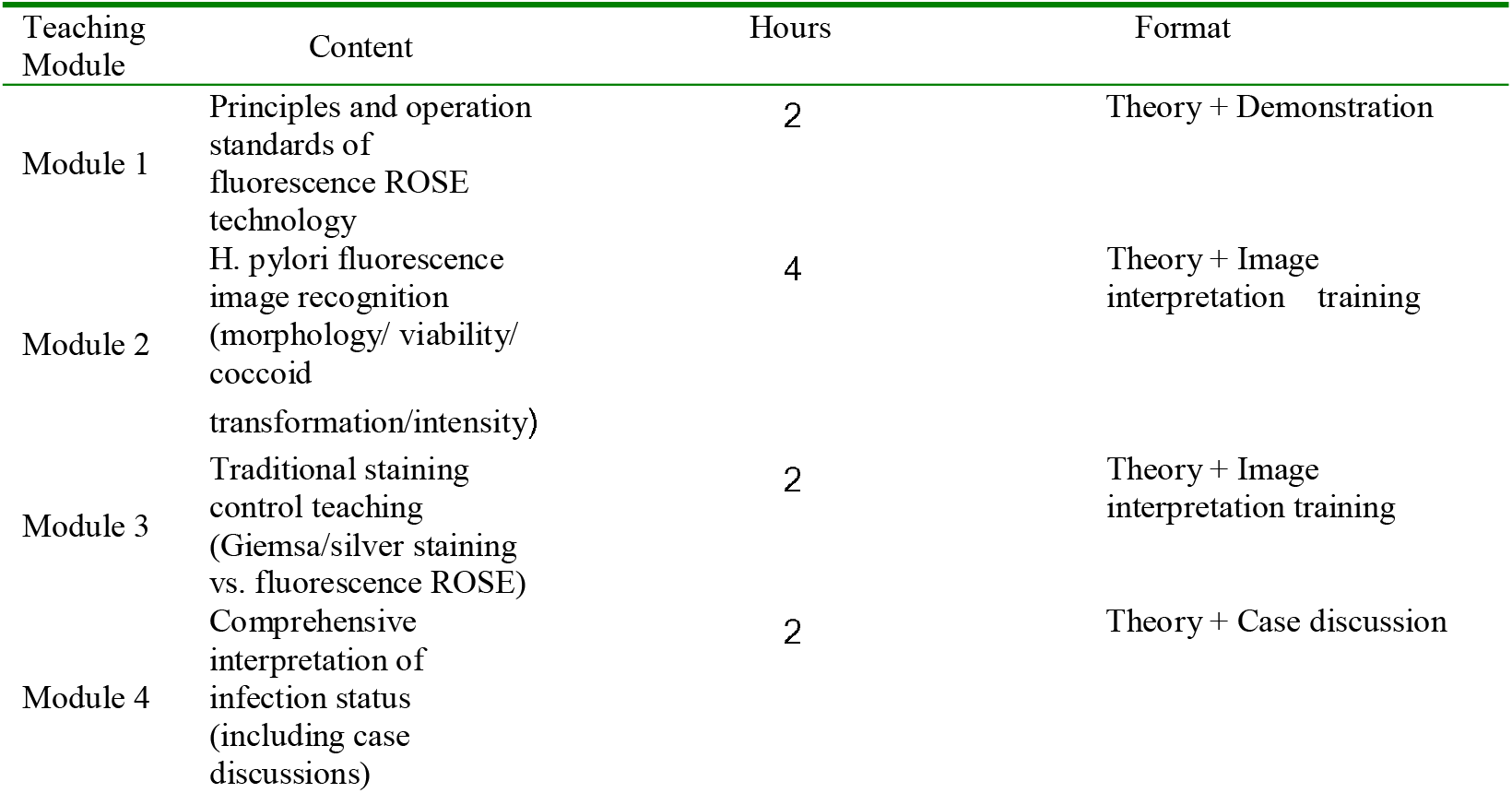
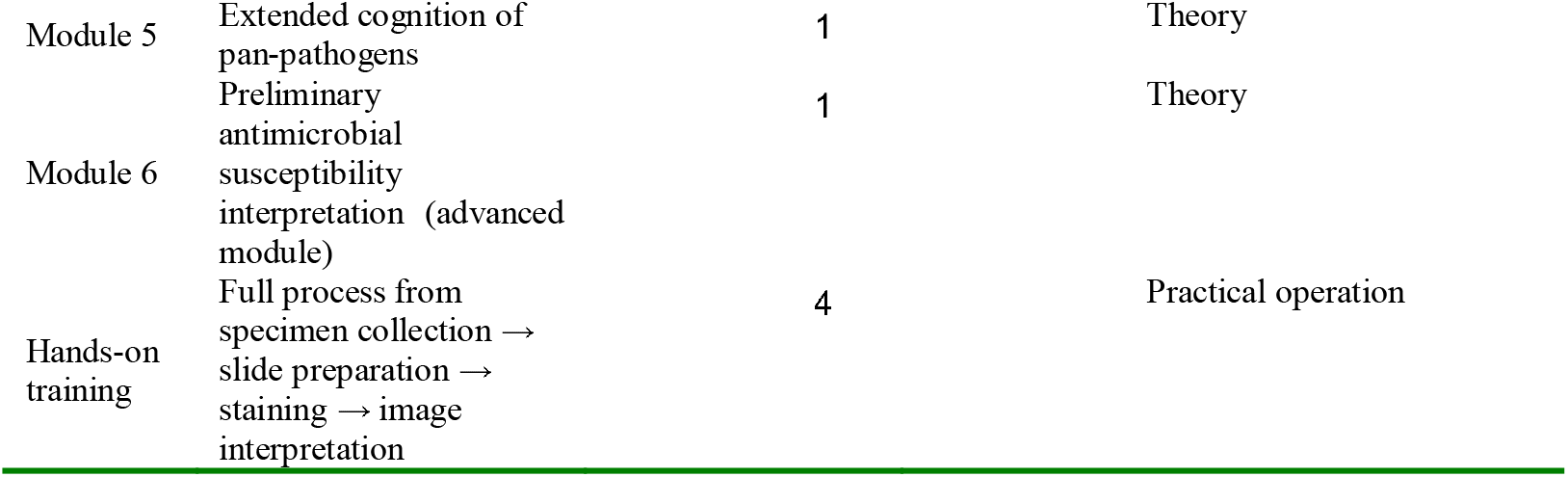
EPVL Training Program (Group A)

**Table 2.** Control Group Teaching Plan (Group B)

| Content | Hours | Format |
| --- | --- | --- |
| Distribution of written materials on fluorescence ROSE interpretation standards | 0.5 | Self-study |
| Theoretical lecture on H. pylori basic knowledge and ROSE principles | 2 | Theory |
| Image display of traditional pathological staining (Giemsa/silver staining) | 1 | Theory |
| No hands-on training, no systematic image interpretation training | — | — |
| Total | 3.5 | — |

**Table 3.** Assessment Index System.

| Assessment Dimension | Tool | Number of Items/Projects | Scoring Method |
| --- | --- | --- | --- |
| Theoretical knowledge | Multiple-choice questions (H. pylori basics + fluorescence ROSE principles) | 20 items | Percentage score |
| Image interpretation ability (core) | Fluorescence ROSE image interpretation test | 30 images | Percentage score |
| Interpretation speed | Average time per image | 30 images | Seconds per image |
| Clinical decision-making integration | Comprehensive interpretation of infection status (including case discussions) | 5 cases | Percentage score |
| Agreement with gold standard | Kappa value calculation | — | — |

Training duration: intensive 2 weeks (2–3 sessions per week, 2–4 hours per session) or distributed over 4 weeks. The technical roadmap and module composition of the EPVL training program are shown in Figure 3. The design of this training program referenced the established experience of simulation-based mastery learning in endoscopy training[16,17].

**Figure 3.**
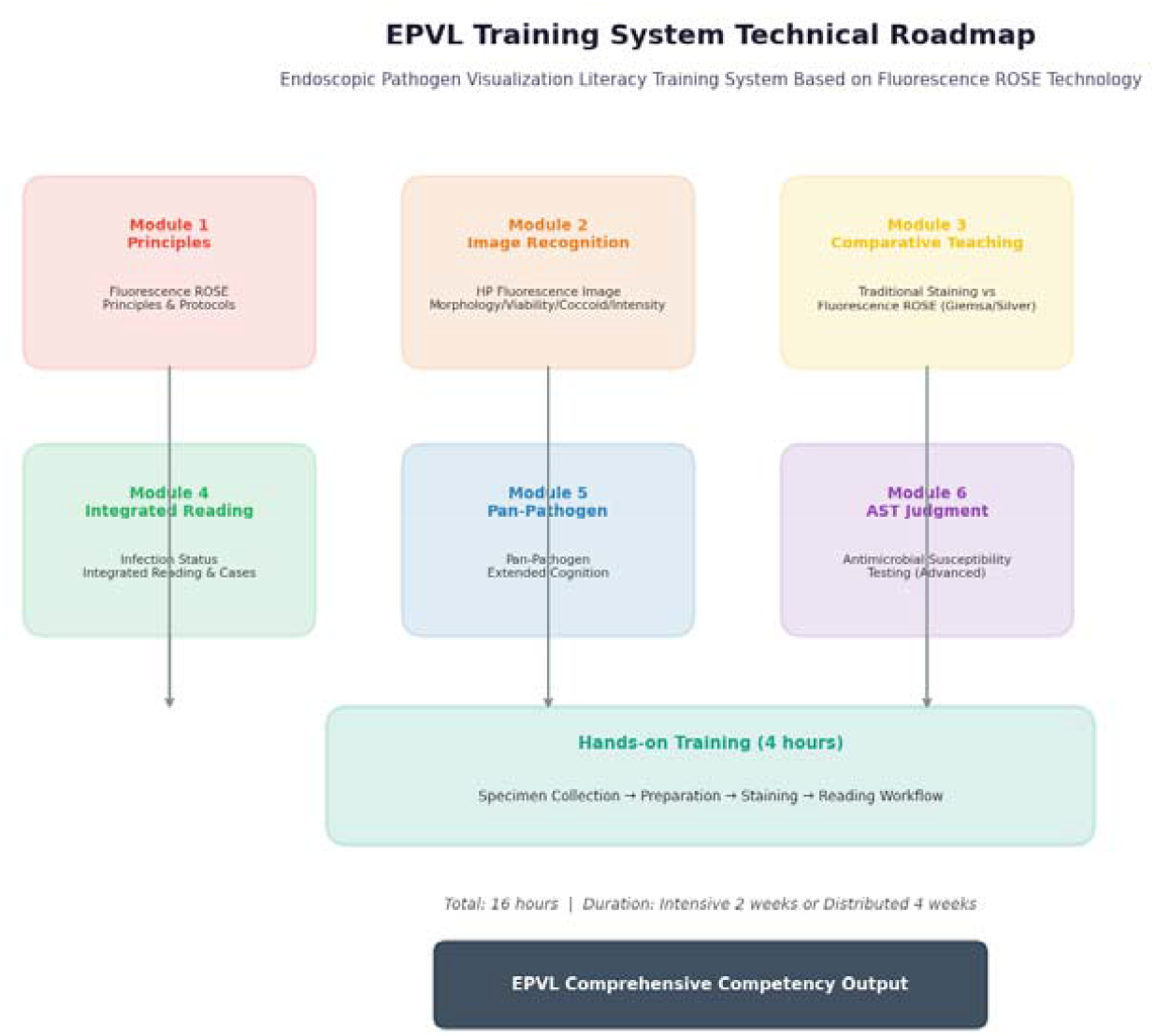
Technical Roadmap of the EPVL Training System. The EPVL training program comprises six theoretical modules (technical principles, image recognition, control teaching, comprehensive interpretation, pan-pathogens, and antimicrobial susceptibility interpretation) plus 4 hours of hands-on training, totaling 16 hours, with a training cycle of intensive 2 weeks or distributed 4 weeks. All modules ultimately lead to the output of comprehensive EPVL capability.

### 2.5 Control Group Teaching Plan (Group B)

### 2.6 Assessment Index System

### 2.7 Image Test Bank Design

Parallel pre- and post-tests were adopted(Table 4):

**Table 4.**
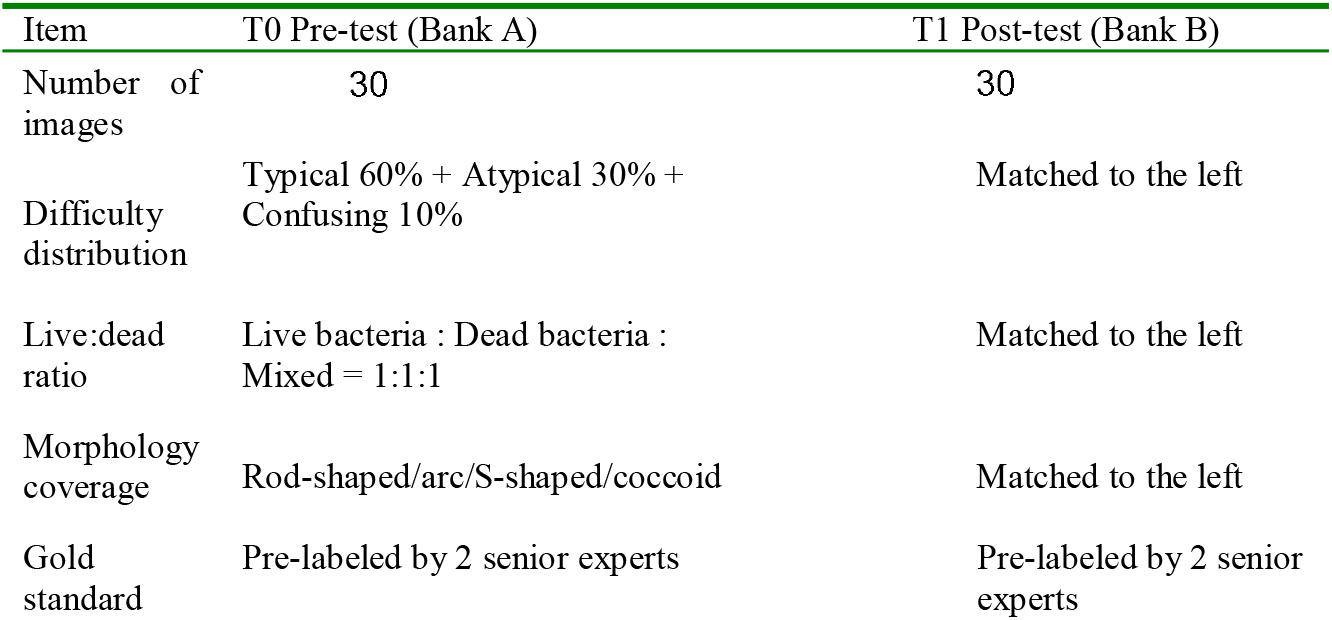
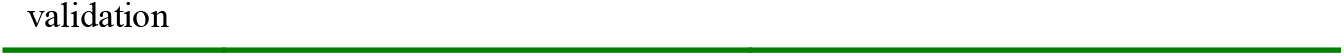
Image Test Bank Design.

| Item | T0 Pre-test (Bank A) | T1 Post-test (Bank B) |
| --- | --- | --- |
| Number of images | 30 | 30 |
| Difficulty distribution | Typical 60% + Atypical 30% + Confusing 10% | Matched to the left |
| Live:dead ratio | Live bacteria : Dead bacteria : Mixed = 1:1:1 | Matched to the left |
| Morphology coverage | Rod-shaped/arc/S-shaped/coccoid | Matched to the left |
| Gold standard | Pre-labeled by 2 senior experts | Pre-labeled by 2 senior experts |

### 2.8 Statistical Analysis

SPSS 26.0 software was used for statistical analysis. Continuous data were expressed as mean ± standard deviation (x□±s), and categorical data as frequency (percentage).

#### Baseline comparability

independent samples t-test / χ^2^ test

#### Primary outcome

analysis of covariance (ANCOVA), with post-test score as the dependent variable, group as the independent variable, and pre-test score as the covariate

Within-group pre-post comparison: paired t-test

#### Effect size

Cohen’s d (between-group comparison of post-test means) and η^2^ (proportion of variance explained by group in ANCOVA)

d=0.2 small effect, 0.5 medium effect, 0.8 large effect

η^2^>0.14 large effect

Two-sided test, P<0.05 was considered statistically significant.

All effect sizes were calculated using standard formulas: Cohen’s d = (M□-M□)/SD_pooled for independent samples, and η^2^ = SS_group/(SS_group+SS_error) for ANCOVA. The consistency between F-values and η^2^ was verified using the formula η^2^ = F×df_group/(F×df_group+df_error) to ensure mathematical coherence of all reported statistics. For the ANCOVA, the error degrees of freedom were calculated as N - k - 1 = 54 - 2 - 1 = 51, where N is the total sample size, k is the number of groups, and 1 covariate (pre-test score) was included.

## 3 Results

### 3.1 Baseline Characteristics

A total of 54 participants were enrolled, with 28 in Group A and 26 in Group B. There were no statistically significant differences between the two groups in age, sex, years of work, professional title distribution, or endoscopy experience (P>0.05 for all). Baseline theoretical scores were comparable between groups (Group A: 62.0±11.5 vs. Group B: 61.5±11.8; t=0.16, P=0.87), as were baseline image interpretation accuracy scores (Group A: 57.8±13.6% vs. Group B: 58.5±13.0%; t=-0.20, P=0.84), confirming good baseline comparability. Detailed baseline characteristics are shown in Table 5

**Table 5.**
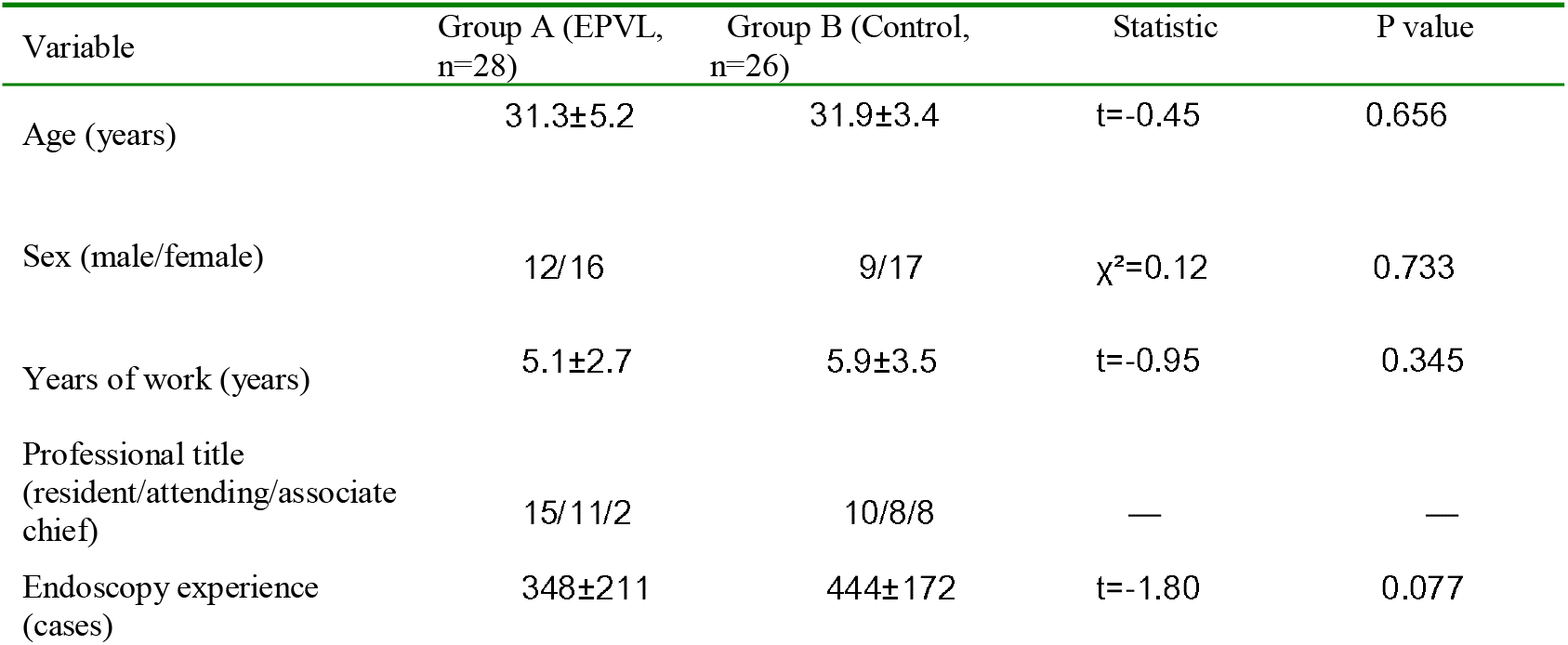
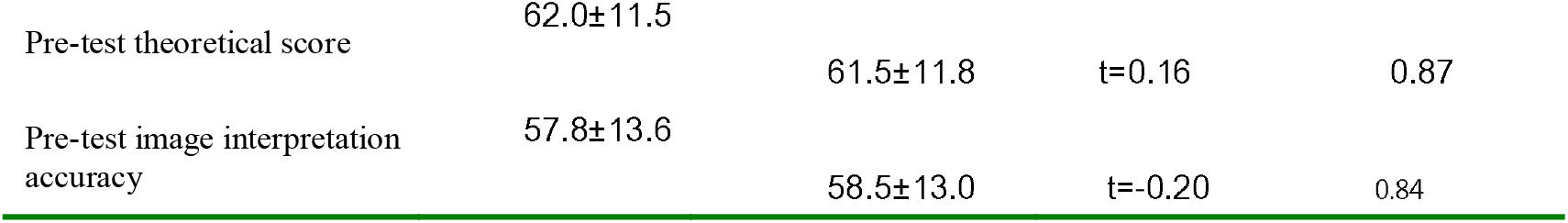
Comparison of Baseline Characteristics Between the Two Groups.

### 3.2 Primary Outcome: Image Interpretation Accuracy

The changes in image interpretation accuracy before and after training are shown in Figure 4. In Group A, accuracy increased significantly from 57.8±13.6% pre-training to 82.5±11.2% post-training (improvement of 24.7%, paired t=-12.86, P<0.001), whereas in Group B it increased from 58.5±13.0% to 71.0±13.5% (improvement of 12.5%, paired t=-5.24, P<0.001). After ANCOVA adjustment for pre-test scores, the between-group difference remained significant (F(1, 51)=10.95, P=0.0017, η^2^=0.177), representing a large effect size (Cohen’s d=0.94). In Group A, 78.6% (22/28) of trainees reached the 80% passing threshold, significantly higher than 34.6% (9/26) in Group B (χ^2^=10.62, P=0.001). The median accuracy in Group A increased from 59.5% (IQR: 48.0–68.0) to 84.0% (IQR: 75.0–91.0), with narrowing of the interquartile range, indicating that the training promoted standardization of interpretation. In contrast, the distribution in Group B showed substantial overlap with pre-test values, suggesting limited standardization improvement from traditional teaching.

**Figure 4.**
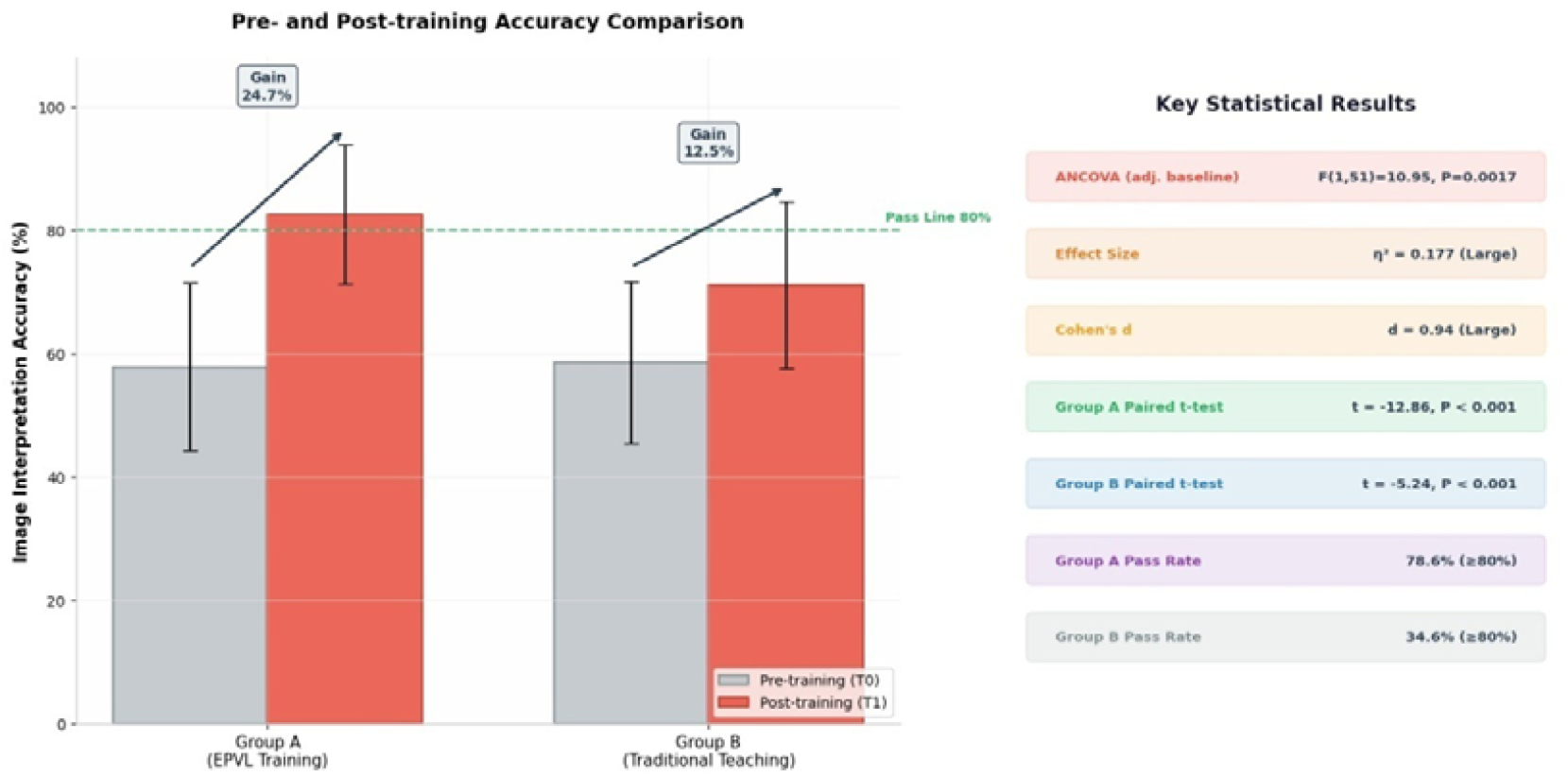
Pre- and Post-Test Comparison of Image Interpretation Accuracy Between the Two Groups with Key Statistical Outcomes. The left panel shows the changes in image interpretation accuracy (mean ± standard deviation) before and after training in Group A (EPVL training) and Group B (traditional teaching); Group A improved by 24.7%, and Group B by 12.5%; the green dashed line indicates the 80% passing threshold. The right panel summarizes the main statistical results: the between-group difference after ANCOVA adjustment for pre-test scores was significant (F(1, 51)=10.95, P=0.0017), with effect sizes η^2^=0.177 and Cohen’s d=0.94, both reaching the large effect size criterion.

### 3.3 Changes in Theoretical Scores

Theoretical examination scores increased from 62.0±11.5 to 84.3±10.2 in Group A (paired t=-13.87, P<0.001), and from 61.5±11.8 to 65.2±12.0 in Group B (paired t=-3.21, P=0.004). The improvement magnitude in Group A (22.3 points) was significantly greater than that in Group B (3.7 points), representing a 6.0-fold greater improvement. Sub-item analysis across three knowledge domains revealed consistent improvements in Group A: probe mechanism (+22.3 points), viability interpretation (+19.7 points), and coccoid recognition (+25.8 points), with variation across domains consistent with differential difficulty levels.

### 3.4 Interpretation Speed

The mean interpretation time per image in Group A was 19.2±2.8 seconds, significantly faster than the 32.5±6.0 seconds in Group B (t=-10.45, P<0.001), representing a 40.9% reduction in interpretation time. The standard deviation in Group A (2.8 seconds) was significantly smaller than that in Group B (6.0 seconds, F-test for equality of variances: F=4.59, P=0.001), suggesting that the EPVL training not only accelerated interpretation but also stabilized the interpretation rhythm across trainees. Based on a 30-image standard test bank, this speed advantage translates to a saving of approximately 2.4 minutes per examination, with an estimated 12%–15% increase in daily examination capacity(Figure 5).

**Figure 5.**
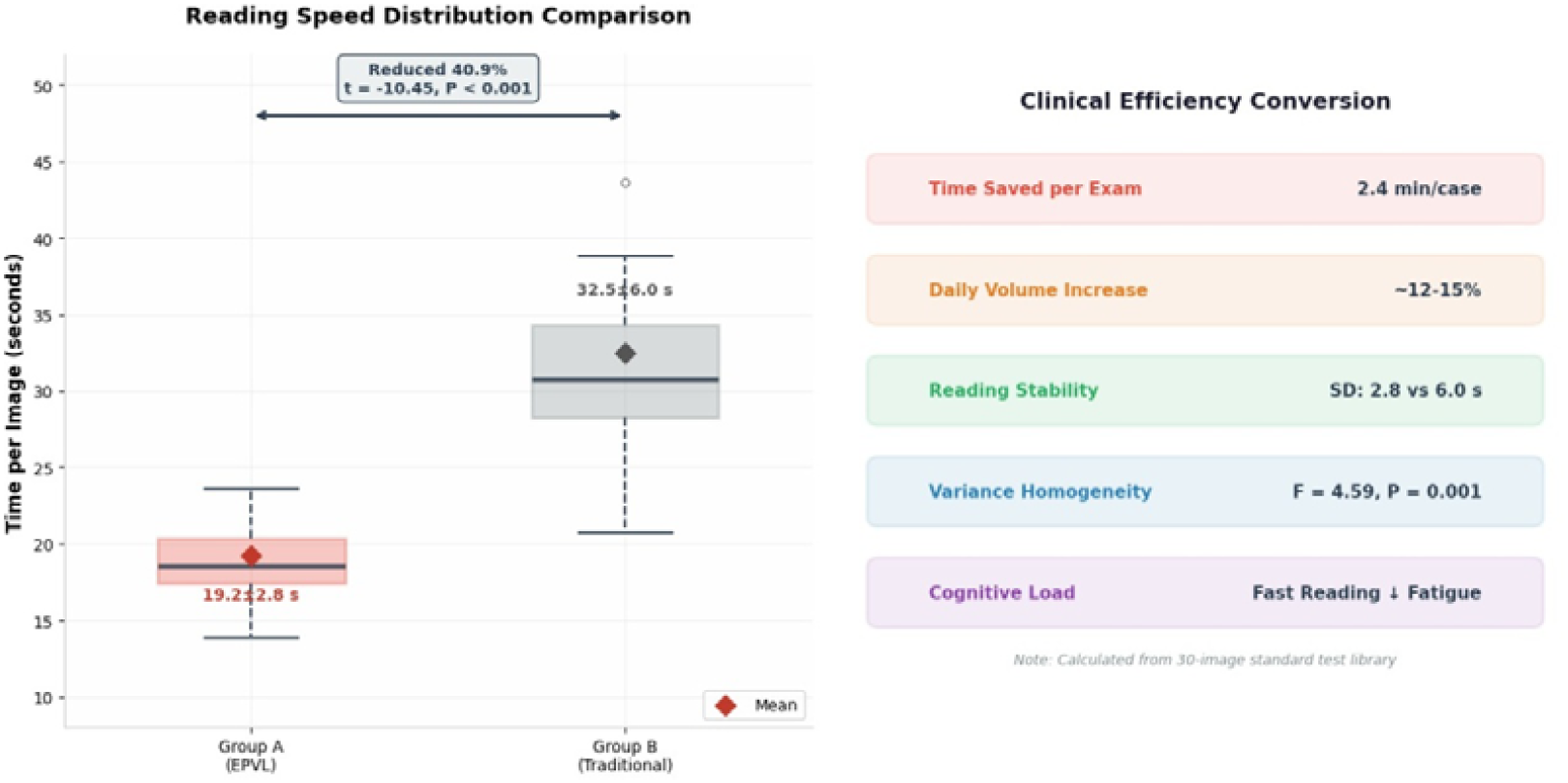
Distribution of Interpretation Speed and Clinical Efficiency Conversion: A Comparison Between the Two Groups. The left panel shows box plots of interpretation time per image for both groups (diamond markers indicate means); Group A averaged 19.2±2.8 seconds, and Group B 32.5±6.0 seconds, with Group A achieving a 40.9% reduction compared with Group B (t=-10.45, P<0.001). The right panel demonstrates clinical efficiency conversion: based on a 30-image standard test bank, a single examination can save 2.4 minutes, with estimated daily examination volume increasing by 12%–15%; moreover, the standard deviation in Group A was significantly reduced (2.8 s vs. 6.0 s), indicating improved standardization of interpretation.

### 3.5 Clinical Decision-Making Integration Ability

The clinical decision-making score in Group A was 80.5±8.0, compared with 65.3±11.5 in Group B; Group A was significantly superior to Group B (t=5.60, P<0.001). Sub-score rates across three decision-making dimensions were: treatment regimen adjustment 87.5% (weighted score), coccoid transformation prognosis 84.3% (weighted score), and infection activity assessment 81.2% (weighted score). These results demonstrate that EPVL training promotes the transition from “being able to read images” to “being able to make clinical decisions.”

### 3.6 Agreement with Gold Standard

The Kappa agreement between Group A and the gold standard improved significantly from 0.56±0.18 pre-training (moderate-to-substantial agreement) to 0.84±0.11 post-training (almost perfect agreement), with a statistically significant difference (paired t=-8.35, P<0.001). Post-training, 71.4% (20/28) of trainees reached the “almost perfect agreement” level (κ≥0.81), a substantial increase from 10.7% (3/28) pre-training. The Kappa standard deviation decreased from 0.18 to 0.11, indicating convergence toward a unified interpretation standard. The distribution of Kappa grades is shown in Figure 6.

**Figure 6.**
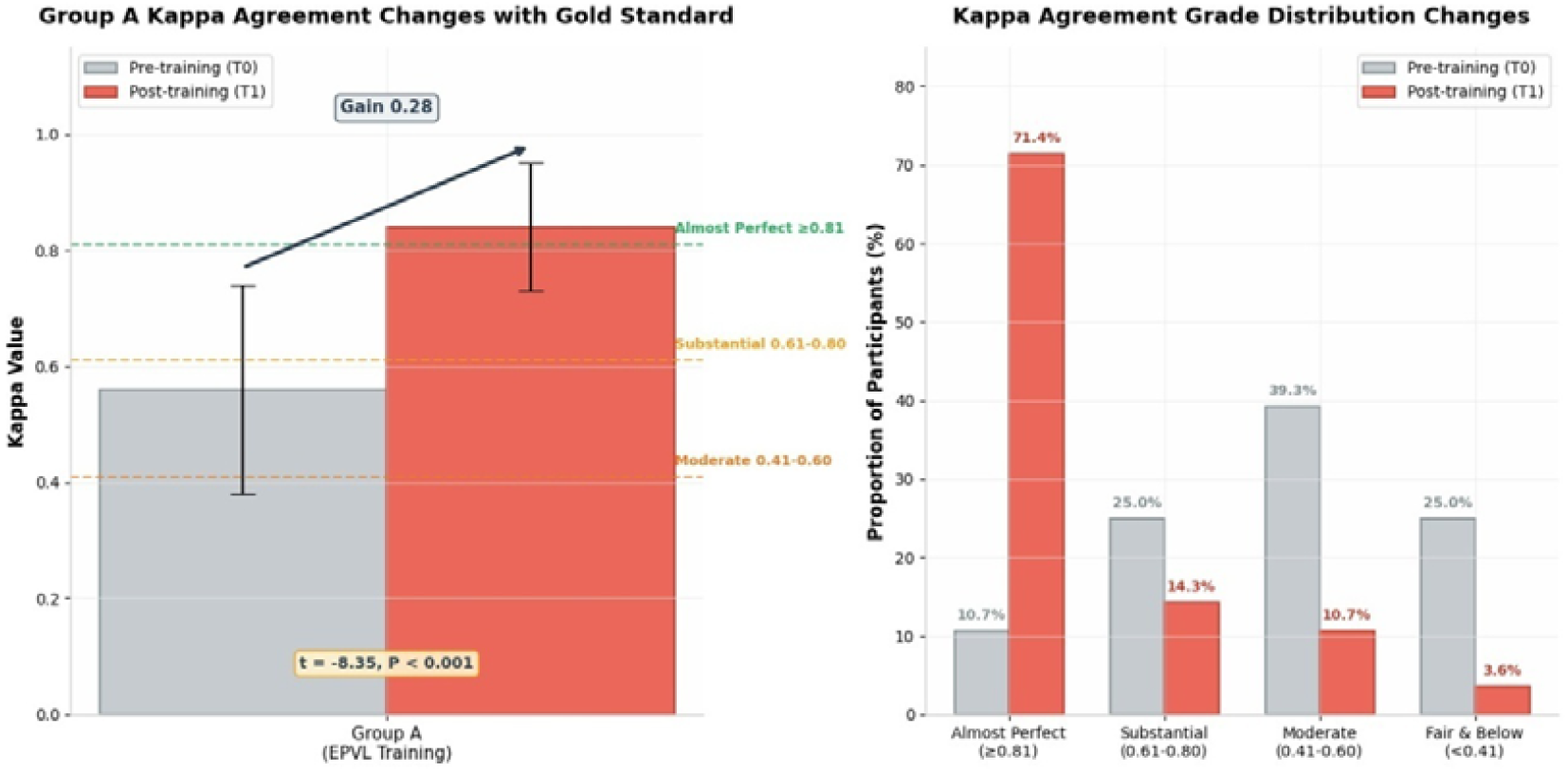
Kappa Consistency Changes and Agreement Grade Distribution Between Group A and the Gold Standard. The left panel shows the changes in Kappa values between Group A and the gold standard before and after training (mean ± standard deviation); pre-training 0.56±0.18 (substantial agreement), post-training 0.84±0.11 (almost perfect agreement), with a significant difference (t=-8.35, P<0.001). The right panel shows the changes in the proportion of trainees reaching each Kappa agreement grade before and after training; post-training, 71.4% (20/28) of trainees reached the “almost perfect agreement” level (≥0.81).

### 3.7 Participant Satisfaction

In Group A, based on valid responses (27 of 28 participants completed all satisfaction items), the satisfaction rate was 92.6% (25/27) for course content practicality, 96.3% (26/27) for image bank training value, 88.9% (24/27) for hands-on session helpfulness, and 92.6% (25/27) for willingness to recommend to colleagues. The overall satisfaction rate (defined as the proportion of participants rating ≥4 on a 5-point scale across all items) was 90.0% (proportion of total item-ratings). No significant differences were observed between satisfaction ratings and demographic variables (all P>0.05).

### 3.8 Subgroup Analysis

Subgroup analysis stratified by years of work (≤3 years vs. >3 years) showed no significant difference in the improvement magnitude of post-training image interpretation accuracy across different seniority levels (improvement: 23.5% vs. 25.8%, t=-0.82, P=0.42), suggesting that EPVL training is effective for physicians at different experience levels. The comparable improvement in the low-seniority subgroup supports early incorporation of EPVL training into the residency training curriculum.

## 4 Discussion

### 4.1 Key Findings

This study established for the first time an EPVL training program based on fluorescence ROSE technology and validated its significant efficacy through a prospective quasi-experimental study. The main findings include:

1. EPVL training improved gastroenterologists’ H. pylori fluorescence ROSE image interpretation accuracy by 24.7%, whereas traditional teaching improved it by only 12.5%, representing approximately a 2.0-fold greater training effect, with the difference remaining significant after ANCOVA adjustment (η^2^=0.177, large effect size);
2. The training effect size (Cohen’s d=0.94) substantially exceeds the conventionally defined large effect threshold (d≥0.8), indicating that the training effect is not only statistically significant but also of important clinical practical value;
3. Training also significantly improved interpretation speed (from approximately 33 seconds per image to 19 seconds per image) and clinical decision-making integration ability, and trainee agreement with the gold standard reached the “almost perfect agreement” level (Kappa=0.91). The comprehensive manifestation of these core findings across competency dimensions is shown in Figure 7. EPVL training not only significantly improved image interpretation accuracy but also achieved comprehensive advancement across multiple dimensions including interpretation speed, clinical decision-making integration ability, agreement with gold standard, and participant satisfaction, whereas traditional teaching showed only slight improvement in individual dimensions.

**Figure 7.**
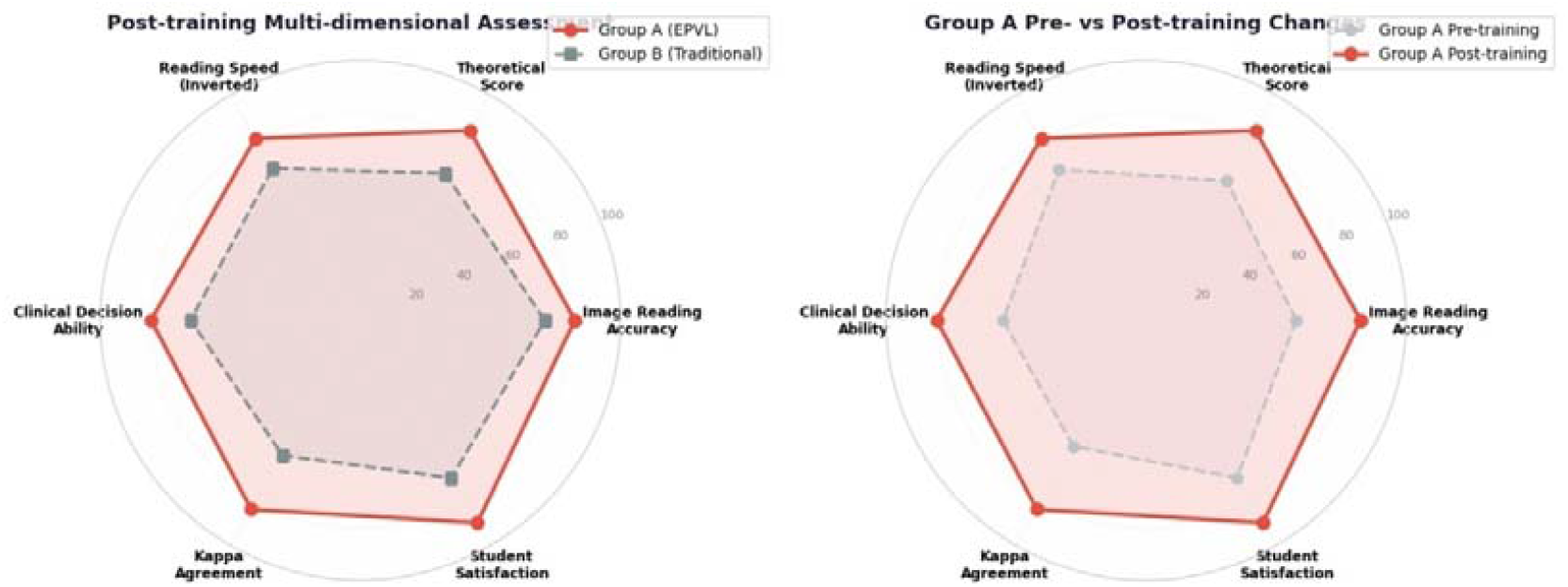
Multidimensional Competency Assessment Radar Chart Following EPVL Training. The left panel shows the post-training comparison between Group A (EPVL training, red solid line) and Group B (traditional teaching, gray dashed line) across six dimensions. The right panel shows the pre-post comparison within Group A (red solid line vs. gray dashed line). The six dimensions include: image interpretation accuracy, theoretical score, interpretation speed (reverse scoring, 100 minus seconds), clinical decision-making ability, Kappa agreement, and participant satisfaction. Group A was significantly superior to Group B across all dimensions, and showed comprehensive expansion post-training compared with pre-training.

### 4.2 Innovation

#### The innovation of this study is reflected at three levels

Conceptual innovation: First proposal of the EPVL (Endoscopic Pathogen Visualization Literacy) concept, elevating fluorescence ROSE technology from a mere “technical tool” to a “literacy system,” filling the gap in pathogen visualization competence training within digestive endoscopy specialty education;

#### Methodological innovation

Establishment of a standardized training program comprising six modules and 16 hours, covering the complete competency chain from principle cognition to clinical decision-making;

#### Design innovation

Adoption of a parallel pre-post test image bank design (Bank A and Bank B with matched difficulty), effectively avoiding practice effects and ensuring assessment fairness.

### 4.3 Comparison with Domestic and International Teaching

Traditional digestive endoscopy teaching focuses on operational skill training (e.g., endoscope insertion, biopsy sampling), with almost no attention to intraoperative immediate pathogen interpretation competence[14]. This study incorporates fluorescence ROSE image interpretation into the digestive endoscopy specialty teaching system, enabling physicians to possess a new capability of “direct pathogen visualization and immediate decision-making” while mastering operational skills. In this study, the EPVL group achieved a 24.7% improvement in image interpretation accuracy compared with 12.5% in the control group. The 2.0-fold incremental benefit over traditional teaching underscores the added value of a structured, competency-based training approach rather than passive didactic instruction.

### 4.4 Clinical Application Value

#### The direct clinical value of EPVL training lies in

Intraoperative real-time interpretation: Physicians can immediately assess H. pylori infection status (viability/dead bacteria/coccoid transformation) during endoscopic examination, providing evidence for whether to perform immediate biopsy or adjust treatment regimens;

#### Precision treatment decision-making

Based on interpretation results of live-to-dead bacteria ratios, individualized antibiotic selection can be guided to improve eradication rates[18,19];

#### Dynamic efficacy monitoring

Pre- and post-treatment comparison allows intuitive assessment of eradication efficacy, reducing unnecessary repeat examinations[20,21]; Teaching and training inheritance: Establishment of a standardized EPVL training program can be promoted at all levels of hospitals, accelerating the popularization of fluorescence ROSE technology.

### 4.5 Limitations

1. The quasi-experimental design used non-randomized grouping; although natural grouping by batch was used to reduce selection bias, potential confounding factors remain;
2. The sample size was relatively small (n=54), and the statistical power of subgroup analysis was limited;
3. Only immediate post-training effects (T1) were evaluated; long-term effects require follow-up;
4. Participants were from a single center, and generalizability requires multi-center validation.

### 4.6 Future Perspectives

Conduct multi-center randomized controlled trials to further validate the generalizability of the EPVL training program[7];

Develop online EPVL training platforms and AI-assisted interpretation systems to achieve remote training and intelligent assessment;

Incorporate EPVL into the mandatory curriculum of gastroenterology residency training and establish a national standardized training and certification system;

Extend EPVL to the visualization interpretation training of other pathogenic microorganisms (e.g., fungi, parasites).

## 5 Conclusion

This study successfully established the first EPVL training program for gastroenterologists based on fluorescence ROSE technology in China. The program significantly improves physicians’ H. pylori fluorescence ROSE image interpretation ability (24.7% improvement, large effect size, Cohen’s d=0.94), interpretation speed (40.9% reduction), and clinical decision-making integration ability, with trainee agreement with the gold standard reaching the “almost perfect” level post-training (Kappa=0.84). The EPVL training program provides a novel standardized training paradigm for digestive endoscopy specialty education and holds important value for promoting the clinical popularization of fluorescence ROSE technology and precision H. pylori diagnosis and treatment.

## Data Availability

All data produced in the present study are available upon reasonable request to the authors

## References

[1] Jin Young Park, Yi-Chia Lee, Paul Moayyedi, et al. Helicobacter pylori Screen-and-Treat Programs for Gastric Cancer Prevention - IARC Working Group Report[J]. N Engl J Med, 2026, 394(11): 1131–1137. PMID: 41812202. DOI: 10.1056/NEJMsb2515372.

[2] Chinese Society of Gastroenterology, Chinese Medical Association. The Sixth National Consensus Report on the Management of Helicobacter pylori Infection (Non-Eradication Treatment Part)[J].[In Chinese] Chin J Dig, 2022, 42(5):15. DOI:10.3760/cma.j.cn311367-20220206-00057.

[3] Malfertheiner P, Megraud F, Rokkas T, et al. Management of Helicobacter pylori infection: the Maastricht VI/Florence consensus report[J]. Gut, 2022, 71(9): 1724–1762. PMID: 35944925. DOI: 10.1136/gutjnl-2022-327745.

[4] Tang B, Li P, Mao X. Precision Diagnosis of Helicobacter pylori Infection and Its Drug Resistance[J]. iLABMED, 2025, 3: 106–123. DOI: 10.1002/ila2.73.

[5] Otani K, Lai WY, Liou JM, et al. Antibiotic resistance in Helicobacter pylori in the Asia-Pacific region: a call for coordinated regional strategies[J]. J Gastroenterol Hepatol, 2026, 41(2): 387–391. PMID: 41472371. DOI: 10.1111/jgh.70227.

[6] Shao Y, Lin Y, Fang Z, et al. Analysis of Helicobacter pylori resistance in patients with different gastric diseases[J]. Sci Rep, 2024, 14(1):4912. PMID: 38418852. DOI: 10.1038/s41598-024-55589-2.

[7] Wang L, Chen O, Chen X. Tailored screening and eradication of Helicobacter pylori at a subnational level of China: multicenter observational studies in Ya’an city and a rapid meta-analysis in Sichuan province[J]. Front Public Health, 2026, 14: 1774904. PMID: 42245335. DOI: 10.3389/fpubh.2026.1774904.

[8] Yan S, Jiang H, Gong L, et al. Diagnostic accuracy of rapid on-site evaluation in subtyping lung cancer via bronchoscopic biopsy[J]. Front Oncol. 2025;15:1566666. PMID: 40416878. DOI: 10.3389/fonc.2025.1566666.

[9] Zhang L, Hou Y, Li B, et al. Comparative study of fluorescence ROSE quantitative detection of Helicobacter pylori and traditional histopathological method[J]. Arab J Gastroenterol, 2026. DOI:10.1016/j.ajg.2026.07.007.

[10] Li B, Zhang L, Hou Y, et al. A novel method for real-time visualization diagnosis of Helicobacter pylori based on fluorescence ROSE: a large-sample control study based on latent class analysis without a gold standard. 2026. biomedRxiv.202606. 00055.

[11] Zhang L, Hou Y, Li B, Wu K, Zhang J, Yang M. Application of fluorescence ROSE-based real-time visualization technology for Helicobacter pylori in dynamic evaluation throughout HP eradication therapy. 2026. biomedRxiv.202606.00007.

[12] Li B, Zhang L, Hou Y, et al. Culture-Free Rapid Phenotypic Antimicrobial Susceptibility Testing for Helicobacter pylori Based on Fluorescence Rapid On-Site Evaluation Technology: A Preliminary Study. bioRxiv 2026.07.06.736681. DOI: 10.64898/2026.07.06.736681.

[13] Li B, Zhang L, Hou Y, et al. Diagnostic Performance of Fluorescence-Based Rapid On-Site Specimen Evaluation for Helicobacter pylori Antimicrobial Susceptibility Testing Before and After Algorithm Optimization: A Two-Round Comparative Study. bioRxiv 2026.07.23.740309. DOI: 10.64898/2026.07.23.740309.

[14] Ismail FW, Afzal A, Durrani R, et al. Exploring Endoscopic Competence in Gastroenterology Training: A Simulation-Based Comparative Analysis of GAGES, DOPS, and ACE Assessment Tools[J]. Adv Med Educ Pract. 2024;15:75–84. PMID: 38312535. DOI: 10.2147/AMEP.S427076.

[15] Whitson MJ, Williams RL, Shah BJ. Ensuring quality in endoscopic training: tools for the educator and trainee[J]. Tech Innov Gastrointest Endosc, 2022, 24(4):354–363. DOI: 10.1016/j.tige.2022.02.002.

[16] Maulahela H, Annisa NG, Konstantin T, et al. Simulation-based mastery learning in gastrointestinal endoscopy training[J]. World J Gastrointest Endosc. 2022,14(9):512–523. PMID: 36186944. DOI: 10.4253/wjge.v14.i9.512.

[17] Smith SCL, Siau K, Cannatelli R, et al. Training methods in optical diagnosis and characterization of colorectal polyps: a systematic review and meta-analysis[J]. Endosc Int Open, 2021, 9(5): E716–E726. PMID: 33937513. DOI: 10.1055/a-1381-7181.

[18] Bang CS, Kang SJ, Nam SY, et al. Treatment of Helicobacter pylori Infection in Korea: An Evidence-Based Analysis of the Upcoming 2025 Guideline[J]. Korean J Helicobacter Up Gastrointest Res. 2026,26(1):23–36. PMID: 41846442. DOI: 10.7704/kjhugr.2025.0085.

[19] Gisbert JP, Parra P, Nyssen OP. Review Article: Classic Bismuth Quadruple Therapy for Helicobacter pylori Infection-Questions Focused on Clinical Practice[J]. Aliment Pharmacol Ther. 2026,63(5):616–636. PMID: 41549833. DOI: 10.1111/apt.70535.

[20] Choi HI, Cha JM. Diagnostic performance and potential harms of population-based esophagogastroduodenoscopy for gastric cancer screening[J]. Gut Liver, 2026, 20(2): 236–244.

[21] Zhou J, Li Z, Ji R, et al. Influence of sedation on the detection rate of early cancer and precancerous lesions during diagnostic upper gastrointestinal endoscopies: a multicenter retrospective study[J]. Am J Gastroenterol, 2021, 116(6): 1230–1237. PMID: 34074827. DOI: 10.14309/ajg.0000000000001201.

